# The association between delirium and falls in older adults in the community: a systematic review

**DOI:** 10.1101/2024.03.12.24303708

**Authors:** Charlotte Eost-Telling, Lucy McNally, Yang Yang, Chunhu Shi, Gill Norman, Saima Ahmed, Brenda Poku, Annemarie Money, Helen Hawley-Hague, Chris Todd, Susan D. Shenkin, Emma R.L.C. Vardy

## Abstract

**Objective:** Systematically review and critically appraise evidence for the association between delirium and falls in community-dwelling adults aged 60 years and above

**Methods:** We searched EMBASE, MEDLINE, PsycINFO, Cochrane Database of Systematic Reviews, CINAHL and Evidence-Based Medicine Reviews (EBMR) databases in April 2023. Standard methods were used to screen, extract data, assess risk of bias (using Newcastle Ottawa scale), provide a narrative synthesis and where appropriate conduct meta-analysis.

**Results:** We included eight studies, with at least 3505 unique participants. Five found limited evidence for an association between delirium and subsequent falls: one adjusted study showed an increase in falls (RR 6.66;95% CI 2.16-20.53) but the evidence was low certainty. Four non-adjusted studies found no clear effect. Three studies (one with two subgroups treated separately) found some evidence for an association between falls and subsequent delirium: meta-analysis of three adjusted studies showed an increase in delirium (pooled OR 2.01; 95%CI 1.52-2.66), one subgroup of non-adjusted data found no clear effect. Number of falls and fallers were reported in the studies. Four studies and one subgroup were at high risk of bias and one study had some concerns.

**Conclusions:** We found limited evidence for the association between delirium and falls. More methodologically rigorous research is needed to understand the complex relationship, establish how and why this operates bi-directionally and identify potential modifying factors involved. We recommend the use of standardised assessment measures for delirium and falls. Clinicians should be aware of the potential relationship between these common presentations.

**Key points:**

- This is the first systematic review of the association between delirium and falls in the wider community population.
- There is relatively limited but consistent evidence on the direction of effect for both delirium preceding falls and falls preceding delirium.
- More high-quality longitudinal work is needed to explore the nature of this potentially complex and bidirectional relationship.
- History of falls and delirium should be considered when assessing patients with incidence/suspected incidence of falls or delirium.

## BACKGROUND

Falls, defined as “an unexpected event in which the participants come to rest on the ground, floor, or lower level” [1], affect nearly one-third of community-dwelling adults aged 65 years and older every year. This rises to over 50% for those aged 80 and above [2–4]. While most fall-related injuries are minor, in the UK, over 223,000 falls in people aged 65 and older resulted in hospital admissions between 2021 and 2022 [5]. This has a personal burden in terms of pain, injury, fear of falling, loss of confidence and independence and higher mortality. Falls in the community are estimated to cost the National Health Service (NHS) over £1.7 billion per year [6, 7].

Delirium, is a condition of acute onset, causing altered attention and awareness with an additional disturbance in cognition, which may fluctuate due to an underlying medical cause [8]. The point prevalence of delirium in the community is estimated at between 1–2%, increasing to 14% in people aged over 85 years [9, 10]. In long-term care facilities, the prevalence of delirium among people aged 65 years and over is estimated at between 10–40% [11]. However, it is often under-detected and underdiagnosed in the community and sometimes misdiagnosed as other conditions including dementia, depression and psychosis [12]. Delirium may have considerable burdens in terms of functional or cognitive decline in individuals and the economic burden to the healthcare system due to increased risk of hospitalisation, higher levels of care and institutionalisation [13–15].

Delirium and falls share common risk factors including older age, prior history of falls, impaired balance and gait, visual and auditory impairment, cognitive impairment, and polypharmacy [16]. The relationship between delirium and falls can be complex and bidirectional.

In hospital settings there is an increased incidence of falls in patients with delirium, and increased risk of delirium in people who had falls. A systematic review [17] reported a higher risk of falls for inpatients with delirium than those without delirium across ten studies (median RR 54.5, range 1.4– 12.6). A recent cross-sectional study analysing the association between delirium and falls in a hospital screening program with more than 29,000 patients [18], found those who screened positive for delirium during admission had a significantly increased risk of falling whilst they were an inpatient (adjusted OR 2.81 (95% CI: 2.12, 3.70)). Delirium screening is recommended as a standard part of a falls care pathway [17, 18].

However, little is known about the association between delirium and falls in community settings. Given that falls are a common reason for pre-hospital service use and hospital admission this is important because there is the potential to reduce hospital admissions and resulting healthcare costs [19]. There is no available systematic review considering the relationships between incidence of falls and occurrence of delirium in the community. Our objective was to conduct a rigorous systematic review of the association between delirium and falls in community settings.

## METHODS

We followed Cochrane review methods for systematic reviews and report the review using the Preferred Reporting Items for a Systematic Review and Meta-analysis (PRISMA) guidelines [20]. The review protocol was prospectively registered on PROPSERO (available at http://www.crd.york.ac.uk/PROSPERO/ registration number: CRD42022309982).

### Search Strategy

A literature search, guided by an information specialist, was conducted in April 2023, using search strategies developed around the facets of delirium, falls and older people (for full search strategy see Supplementary material S1). Searches were completed without date restriction and the following databases were searched: EMBASE (Ovid), MEDLINE (Ovid), PsycINFO (Ovid), Cochrane Database of Systematic Reviews (Ovid), CINAHL (EBSCO), Evidence-Based Medicine Reviews (EBMR)(Ovid). Web of Science and Google Scholar were also searched and restricted to the first 200 relevant records returned. Searches were limited to English-language publications. References were checked for additional relevant publications and forward citation searches were conducted on included papers.

### Inclusion and exclusion criteria

We included studies that met the following criteria, (full details in Supplementary material S2).

#### Population

Adults aged 60 years or over, living in community or supported living/residential care settings. Hospital inpatients and people with end-stage disease were excluded. Studies with mixed populations were included only if it was possible to extract the data from community-dwelling older adults as a separate group.

#### Variables

Studies reporting evidence on the association between delirium and falls, regardless of which preceded the other. Studies with any recognised definition of delirium and falls, author definitions, or no definition were accepted.

#### Study design

Observational studies including time-sequence, cohort, and cross-sectional studies. Randomised and non-randomised controlled trials (of any design in relation to allocation) and interrupted time series or controlled before and after studies were included if the intervention and/or control condition was unrelated to falls or delirium prevention.

### Study selection

Search records were imported into Rayyan [21] and duplicate records removed. Two reviewers (LM and BP) independently screened titles and abstracts for relevance and the full texts of potentially relevant studies. Disagreements were resolved through discussion with a third reviewer (CET) if necessary.

### Data Extraction

A predefined data extraction form was used to extract study characteristics, see Box 1. Two reviewers independently extracted data from included studies (LM and YY) and disagreements were resolved through discussion with a third reviewer (CET or EV).

##### Box 1: Data extraction items

- Basic characteristics of studies, including first author, study date, study location, study aim, study design (type, duration, setting), publication type, publication year
- Characteristics of participants, including population type and place of residence, inclusion criteria, the number of participants, mean age of participants, gender of participants and ethnicity of participants
- Variable measurements (assessment tools/method/scales, time points reported) for both delirium and falls
- Temporal association between falls and delirium, i.e., whether falls or delirium were recorded first
- Falls outcomes: number of falls, number of fallers, falls rate per person per year, time to first fall, number of injurious falls
- Delirium outcomes: number of delirium cases detected, number of people with delirium, number of delirium episodes per person, duration, severity and type of delirium (hyperactive, hypoactive or mixed)
- Statistical methods used, and handling of missing data
- Statistical analysis of results

### Quality assessment

Two reviewers (LM and YY) independently assessed the quality of included studies using the Newcastle Ottawa Scale (NOS) [22, 23]. We considered using other tools such as Quality In Prognosis Studies (QUIPS) [22] and Risk Of Bias In Non-randomized Studies of Exposures (ROBINS-E) [24] but they were less appropriate for this review due to the study designs of the included evidence, i.e. studies not set up specifically as prognostic studies or studies of exposure. Disagreements were resolved through discussion or consultation with a third reviewer (CET).

Scores were assigned to studies based on the quality of selection criteria, comparability of groups and outcome (for cohort and cross-sectional) or exposure (for case-control), with a maximum score of 9 for case-control and cohort studies and 8 for cross-sectional studies. The overall score was used to rate the risk of bias, however, studies were also given an overall high risk of bias if any single domain was rated as high risk.

We also considered the certainty of the evidence, guided in this by the GRADE approach for prognostic reviews [25–27] although the nature of the evidence (i.e., not designed as prognostic studies), meant that we did not conduct a full GRADE assessment.

Risk of bias, inconsistency of the results, indirectness of the evidence, imprecision of the statistical analysis and publication bias for each outcome were considered. In assessing imprecision, we accounted for the number of studies contributing to evidence, the size of the studies and width of the confidence intervals in each study.

### Data synthesis

Data analysis was structured by the temporal association between delirium and falls. We report separately on the data from studies where delirium preceded the recorded falls (D-F) and studies where falls preceded a record of delirium (F-D).

We conducted narrative syntheses and present individual studies’ data for outcome measures on forest plots. Where appropriate we conducted meta-analyses in RevMan5.4 [28] using random effects models with the generic inverse variance method.

## RESULTS

### Study selection

After screening 971 records on title and abstract and 21 on full text, eight studies met the inclusion criteria for this review, see Figure 1 for the PRISMA flowchart of the selection process [20].

**Figure 1.**
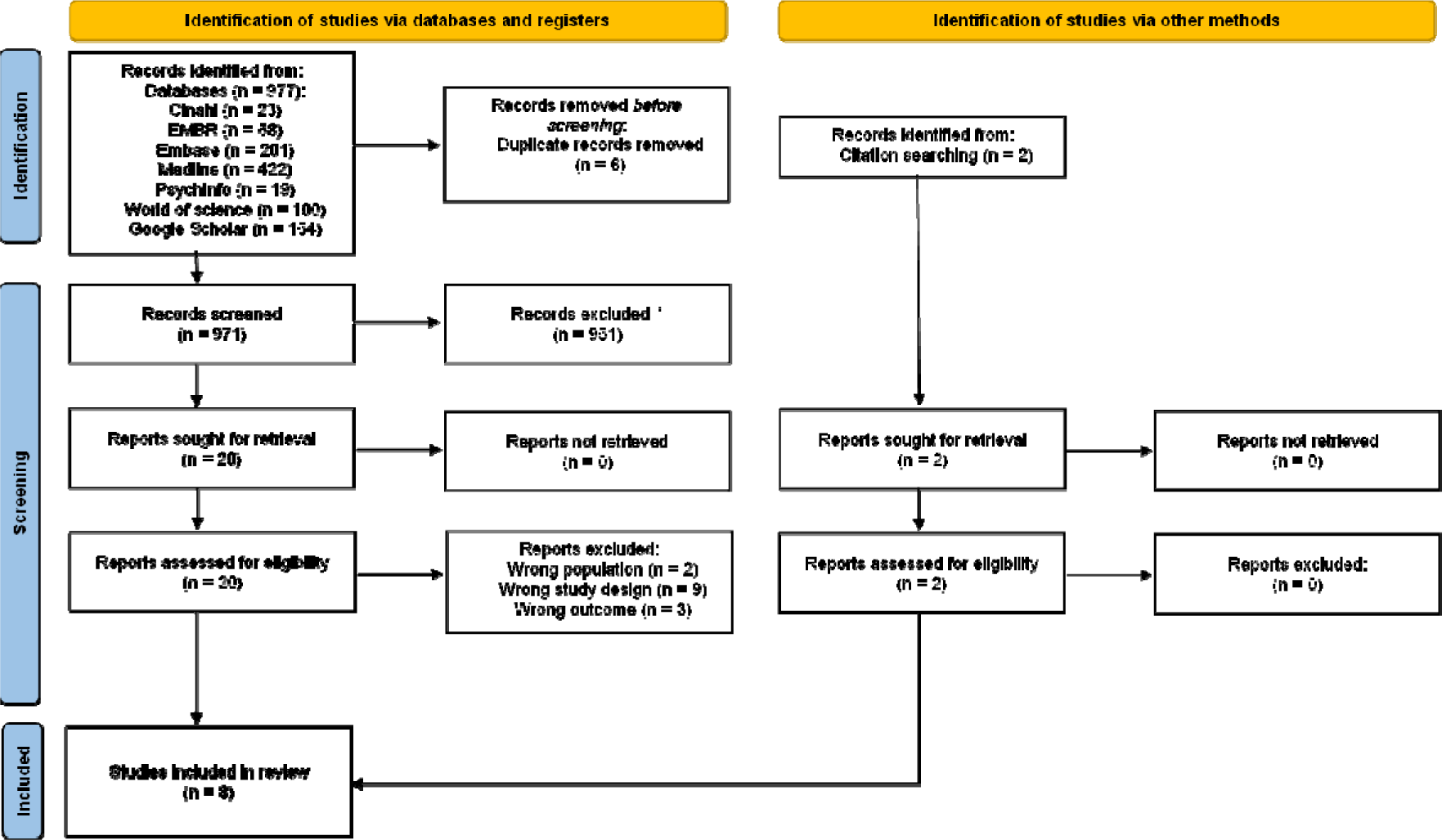
PRISMA Flowchart of study selection process.

### Study and participant characteristics

Eight studies including at least 3505 unique participants from five countries were included in this review. Settings included residential homes and individual homes, and follow-ups ranged from one month to 15.5 months. The main study and participant characteristics are summarised in Table 1 and additional study characteristics can be found in Supplementary S3.

**Table 1:** Study characteristics.

| Delirium-Falls studies |  |  |  |  |  |  |  |  |  |  |  |  |
| --- | --- | --- | --- | --- | --- | --- | --- | --- | --- | --- | --- | --- |
| Study | Study location | Study aim | Study design |  |  |  |  |  |  |  | Variable measurement |  |
| | | | Type | Duration | Study setting | Inclusion criteria | Number of participants | Age (mean years ± SD) | Gender | Ethnicity | Delirium | Falls |
| Eriksson et al. (2007) [29] | Sweden | Identify risk factors for falls in older people with and without a diagnosis of dementia living in residential care facilities (we used only non-dementia subgroup) | Prospective cohort study | 6 month follow up | 4 residential homes | Inclusion:<br>All residents in the care facilities >65yrs<br>Exclusion:<br>Unclear dementia diagnosis<br>Age <65yrs | Older adults without dementia n=83 | 83.5±7.1 years | Male: n=30 (36.1%) | Not reported | Nurse recorded the presence of delirium in past month | Record of falls incidents |
| Kallin et al. (2002) [31] | Sweden | Identification of predisposing and precipitating factors for falls and recurrent falls among frail older people living in residential care facilities | Cross-sectional with prospective follow-up (treated as cohort study) | 12 month follow up | 1 residential home | Inclusion:<br>All residents in the care facility | Older adults n=83 | 79.6±8.6 years | Male: n=25 (30.1%) | Not reported | Medical records for past delirium | Record of falls incidents and recurring falls |
| Study | Study location | Study aim | Study design Type | Duration | Study setting | Inclusion criteria | Number of participants | Age (mean years ± SD) | Gender | Ethnicity | Variable measurement | |
|  |  |  |  |  |  |  |  |  |  |  | Delirium | Falls |
| Kallin et al. (2004) [30] | Sweden | Study precipitating factors for falls among older people living in residential care facilities | Prospective cohort study | 12 month follow up | 5 residential care facilities | Inclusion:<br>All residents in the care facilities | Older adults<br>n=199 | 82.4 ± 6.8 years | Males: n=59 (29.6%) | Not reported | Diagnosed by physician using *DSM-IV* criteria | Record by staff of falls incidents and recurring falls |
| Mahoney et al. (2000) [32] | USA | Evaluate the rate of falls, and associated risk factors, for 90 days following hospital discharge. | Prospective cohort study | 13 weeks | Patient's home | Inclusion<br>Adults ≥65 years discharged from hospital after medical illness, receiving home health care<br>Exclusion<br>In Hospice Care or terminal cancer<br>New CVA or MI in previous 2 months<br>Dementia and no caregiver in home<br>Between hospital discharge and home health care<br>Non-ambulatory<br>Other incl. no phone, unable to speak English | Older adults<br>n=311 | 80.0 ± 7.1 years | Male = 116 (37.2%) | White = 96.8%<br>African American = 2.2% | Confusion Assessment Method (CAM) | Self-reported record of falls on a calendar, with follow-up postcards and telephone interviews |
| Study | Study location | Study aim | Study design Type | Duration | Study setting | Inclusion criteria | Number of participants | Age (mean years ± SD) | Gender | Ethnicity | Variable measurement | |
|  |  |  |  |  |  |  |  |  |  |  | Delirium | Falls |
| von Heideken Wagert et al, (2009) [33] | Sweden | Describe the incidence of falls and fall-related injuries and identify predisposing factors for falls in very old people | Prospective cohort study | 6 month follow up | Ordinary and institutional housing | Inclusion:<br>Residents aged $\geq 85$ years (random sample from half of those over 85yrs and all residents over 90 years) | Older adults n=220 (n=109 in ordinary housing, n=111 institutional housing) | 90.3 ± 4.8 years | Males n=53 (24%) | Not reported | N/R | For those in ordinary housing self-report on a calendar and follow up telephone calls<br>For those in institutional settings, falls reported by staff were reviewed from records |

| Falls-delirium studies |  |  |  |  |  |  |  |  |  |  |  |  |
| --- | --- | --- | --- | --- | --- | --- | --- | --- | --- | --- | --- | --- |
| Study | Study location | Study aim | Study design |  |  |  |  |  |  |  | Variable measurement |  |
| | | | Type | Duration | Study setting | Inclusion criteria | Number of participants | Age (mean years ± SD) | Gender | Ethnicity | Delirium | Falls |
| Boorsma et al. (2012) (Nursing home subgroup) [34] | The Netherlands | Prevalence and incidence of delirium and its risk factors | Retrospective cohort | 4 years 7 months<br>Observation time 10.8 months | 6 nursing homes | Inclusion:<br>• All residents in the nursing home<br>Exclusion:<br>• If observations missed information about delirium | n=828 | No mean age recorded<br>Older age (>85)<br>n=257 (31.0%) | Males n=274 (33.1%) | Not reported | The Nursing Home – Confusion Assessment Method (NH-CAM)[46] | Fall incidents (at least one fall incident in the last 90 days, yes/no) |
| Boorsma et al. (2012) (Residential home subgroup) [34] | The Netherlands | Prevalence and incidence of delirium and its risk factors | Retrospective cohort | 4 years 7 months<br>Observation time 15.5 months | 23 residential homes | Inclusion:<br>• All residents in the residential homes<br>Exclusion:<br>• If observations missed information about delirium | n=1365 | No mean age recorded<br>Older age (>85)<br>n=706 (51.7%) | Males n=346 (25.3%) | Not reported | Positive score on the Nursing Home–Confusion Assessment Method (NH-CAM)[46] | At least one fall incident in the last 90 days yes/no |
| Perez-Ros et al. (2019) [35] | Spain | Identify delirium predisposing and triggering factors and develop a predictive model | Case-control study (relevant data for this review is treated as retrospective cohort) | 12 months | 6 nursing homes | Inclusion:<br>• Residents ≥ 65yrs, living in nursing home during study.<br>Exclusion:<br>• Spent <1yr in the home<br>• Spent >2mo out of the home during study<br>• End-of-life status | n=443 | Mean 85.76± 6.7 years | Males n=96 (21.7%) | Not reported | Geriatrician diagnosed according to DMS-IV criteria and/or Confusion Assessment Method (CAM) | Number of falls during the study period |
| Study | Study location | Study aim | Study design |  |  |  |  |  |  |  | Variable measurement |  |
| | | | Type | Duration | Study setting | Inclusion criteria | Number of participants | Age (mean years ± SD) | Gender | Ethnicity | Delirium | Falls |
| Sabbe et al. (2021) [36] | Belgium | To investigate (point) prevalence of and risk factors for delirium in nursing homes | Cross-sectional (relevant data for this review is treated as a retrospective cohort) | 1 month | 6 nursing homes | Inclusion:<br>Residents of nursing homes ≥ 65yrs<br>Exclusion:<br>In coma<br>Aphasia<br>End-of-life status<br>Specifically on dementia ward | n=338 | Mean 84.7±8.0 years | Males n=110 (32.5%) | Not reported | 13-item Delirium Observation Screening Scale (DOSS) scale based on DSM-IV-TR criteria | Fall incident in previous 90 days - from records |

#### Delirium-Falls (D-F) Studies

Five studies, published between 2000-2009, reported on evidence where recorded delirium preceded falls (D-F) [29–33]. A total of 896 participants were included, with at least 531 unique participants. The additional 365 people could not be confirmed as unique participants (see below).

The five included D-F studies comprised four prospective cohort studies and one follow-up study based on cross-sectional data. Participants were drawn from residential care homes in three studies, their own homes in one study and a mix of residential care and own homes in the final study. Study size ranged from 83 to 311 participants (median 199) and study duration was between 13 weeks and one year (median six months). The median participant age from these five studies was 83.5 years. The proportion of male participants ranged from 21.7% to 37.2%.

Four studies were from Sweden [29–31, 33] and the fifth from the USA [32]. We include evidence from all studies, but this should be interpreted with caution because four studies took place in the same region of Sweden; participants were taken from an ongoing longitudinal study of older people, and it was not clear that participants in each study were unique. We tried to contact the authors of these studies to clarify but did not receive a response. Participants were recruited from the community in four studies [29–31, 33], whilst the USA study enrolled patients in the community after discharge from hospital [32].

Delirium was not explicitly defined in any of the studies. In two studies delirium was identified through patient records, but did not state how it was measured [29, 31]. Two studies measured delirium using either the DSM-IV criteria [30] or the Confusion Assessment Method (CAM) [32]. One study did not state either how delirium had been identified or assessed [33]. Three studies included records of delirium in the previous month [29, 30, 33], one included delirium at baseline [32], and one included historical records of delirium (checked at baseline) [31].

Falls definitions used were mostly variations on standard falls definitions, however only one study provided a referenced definition [32]. Falls were identified either retrospectively from staff records of events or prospectively by self-report on a calendar and a follow-up telephone call from researchers, see Table 1.

#### Falls-Delirium (F-D) Studies

Three studies, one of which stratified data into two subgroups according to type of residence (nursing home or community) and which will be treated as two separate data sets for the purpose of this review, reported evidence when falls preceded recorded delirium (F-D) [34–36]. Studies were published between 2012 and 2021 and included a total of 2974 unique participants.

The three F-D studies included one retrospective cohort, one cross-sectional and one case-control study, all from Europe. However, all the data which were relevant for this review were collected in line with retrospective cohort study design and treated as such for the purposes of this review. Study size ranged from 338 to 1365 participants (median 636) and study duration was between one month and four years seven months (median 12 months). Participants in two studies and one subgroup were recruited from nursing homes and one subgroup was recruited from residential homes.

None of the studies included a description or definition of falls [34–36]. In two studies falls were identified in records from the preceding 90 days [34, 36]. The third study recorded whether a fall had taken place prior to the delirium diagnosis but did not indicate a time frame on this [35].

All three studies described delirium [34–36], but only one provided a referenced definition using the DSM-IV criteria [36]. All studies included details of delirium measures used: two used the CAM [34, 35], one also used criteria from DMS-IV [35] and the final study used the Delirium Observation Screening Scale (DOSS) (based on DSM-IV-TR criteria) [36].

### Quality appraisal

#### Delirium-Falls Studies

Using the appropriate NOS scale for each study, four studies were rated as having a high risk of bias and one study had a low risk of bias (Figure 2 and Supplementary S4a). The highest risk arose because studies did not account for possible confounding factors in the association between delirium and falls in their analysis [29–31, 33].

**Figure 2:**
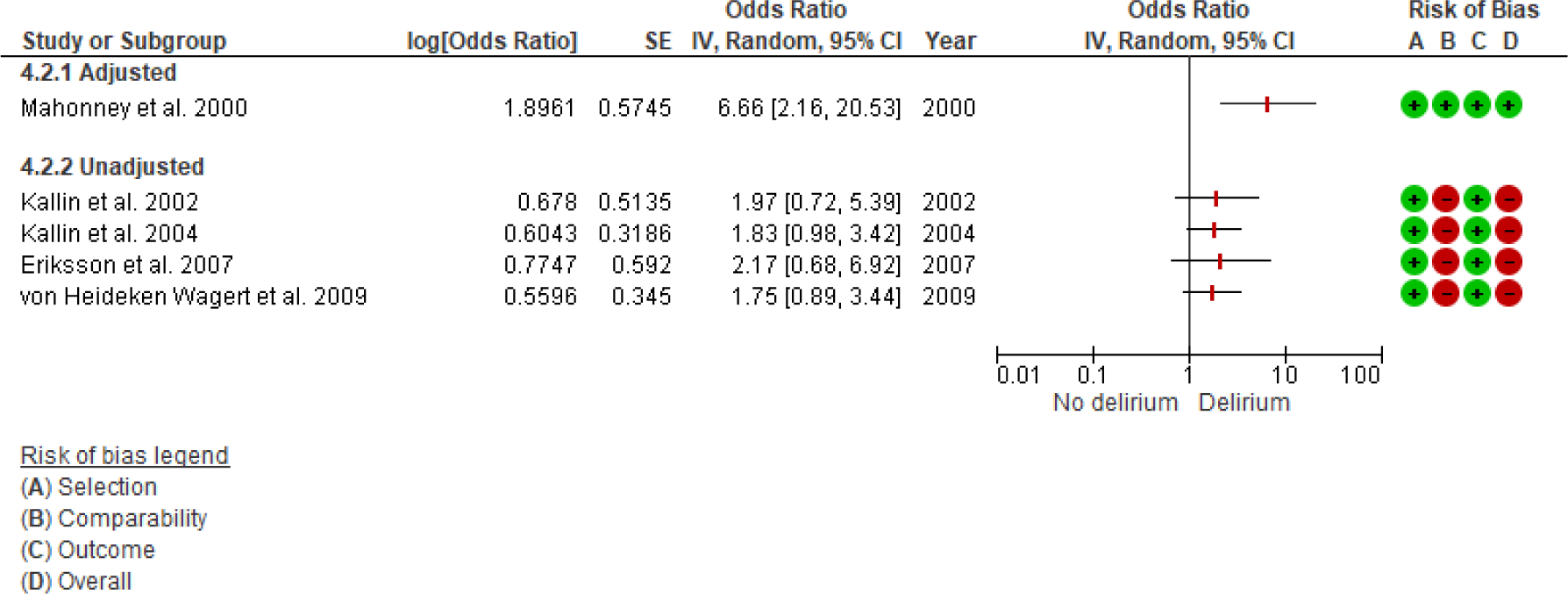
Forest plot of the association between prior delirium and number of falls.

#### Falls-Delirium Studies

One study [35] and one subgroup analysis had a low risk of bias [34], the other subgroup analysis had high risk of bias due to the lack of adjustment for confounding variables relating to falls and delirium [34]. The third study had some concerns over the lack of information provided on participant selection and outcome measures [36] (Figure 3 and Supplementary S4b).

**Figure 3:**
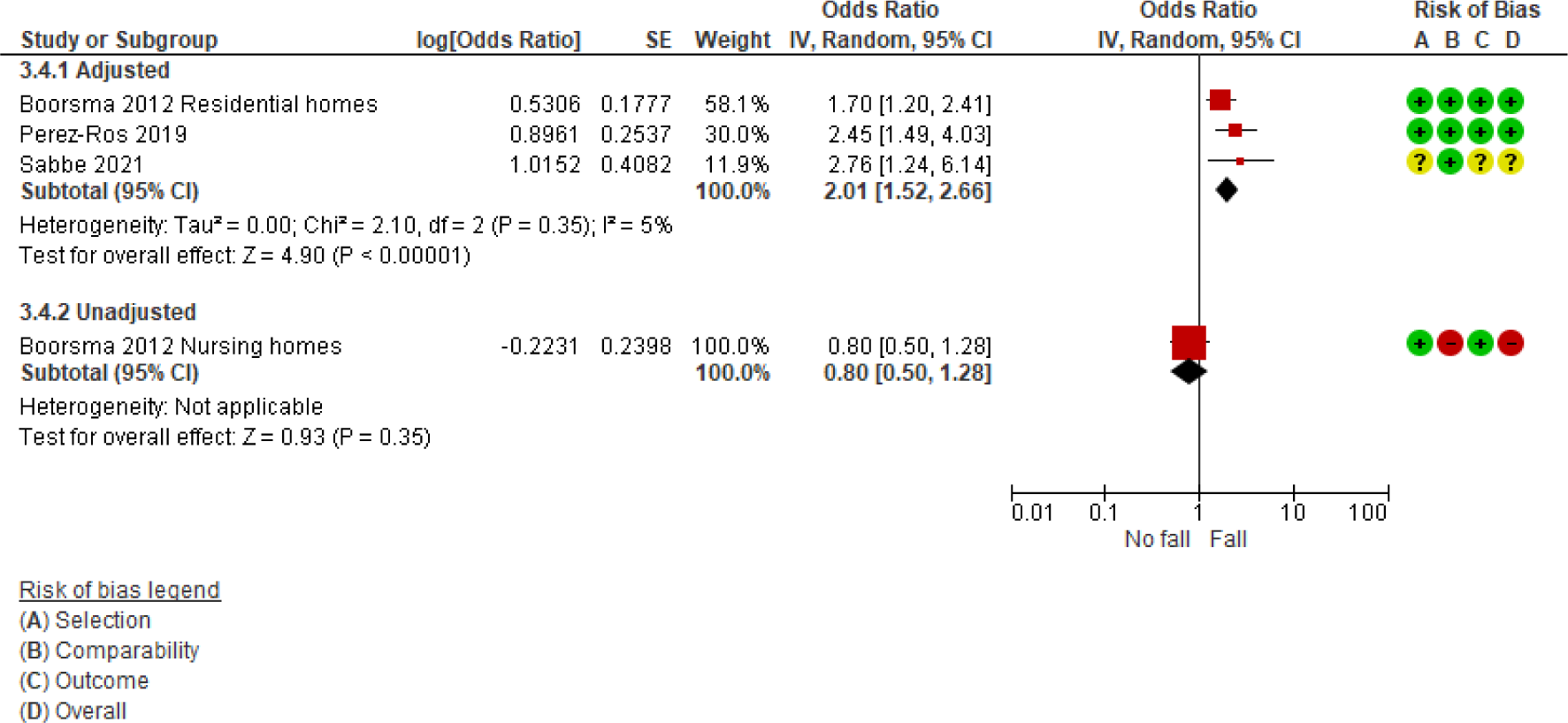
Forest plot of the association between prior falls and delirium.

### Evidence for associations

#### Delirium-Falls Studies

All five studies reported delirium outcomes as the number of people with delirium (five studies [29–33]), none reported the number of delirium episodes per person, duration, severity, or type of delirium. Falls outcomes were reported as the number of falls (four studies [29–31, 33]), number of fallers (five studies [29–33]), falls rate (four studies [29, 31–33]), time to first fall (one study [33]), and number of injurious falls (three studies [30, 32, 33]). Two studies also reported the prevalence of recurrent falls [31, 32]

One study (n=311) presented adjusted data (see Supplementary S5 for details), for the association between delirium and falls [32] and the other four studies (total n=585) presented unadjusted data [29–31, 33]. Meta-analysis was not considered appropriate for the outcomes in these studies because of high heterogeneity in study methodology and analysis, i.e., whether the data was adjusted for confounding variables. All studies are presented in Figure 2 to illustrate effect sizes only.

### Association between delirium and falls

One study (n=311), adjusted for confounding variables, reported a significant association between delirium and an increase in falls in a home setting within 13 weeks of discharge from hospital (RR=6.66, (95% CI 2.16-20.53)), Figure 2 [32]. However, the relevant evidence included in the current review was low certainty due to high imprecision.

The remaining four studies reported unadjusted data on the number of falls, all undertaken within the same regional population. Data across all unadjusted studies was consistent in direction of effect but not statistically significant in any study. The largest and most recently published of these studies (n=220) [33] showed no clear association between delirium and the risk of falls recorded within six months (n=220, RR=1.75 (95% CI, 0.89-1.90)). Evidence from these studies was very low certainty due to the high risk of bias and high imprecision. One non-adjusted study found that participants with a record of delirium in the previous month had a significantly higher risk of falling more than once compared to not falling or only falling once (OR 2.73 [95% CI 1.05, 7.07; p=0.039] [31]. However, this study was considered very low quality, and we have low confidence in the finding.

#### Falls-Delirium Studies

Three studies (one with two subgroups) reported on falls occurring prior to a recorded delirium episode. Falls outcomes reported: number of fallers (three studies [34–36]). No studies reported on whether the fallers experienced single or recurrent falls, the falls rate, time to first fall, or number of injurious falls. Delirium outcomes reported: number of delirium cases (two studies [34, 35]), number of people with delirium (three studies [34–36]), and one study reported on the number of delirium episodes [35]. No studies reported the duration, severity, or type of delirium.

### Association between falls and delirium

Two studies [30, 31] and one study subgroup [34] presented adjusted data on delirium risk (see Supplementary S5 for details). The other study subgroup presented unadjusted data [34]. A random-effects meta-analysis (Figure 3), indicates that the adjusted studies, two at low risk of bias and one with some concerns, showed a significant association between falls and an increased risk of delirium (OR 2.01, 95% CI 1.52-2.66). While the evidence from these studies was graded as high quality, we advise caution. Despite not finding evidence of publication bias, and therefore not downgrading for this, it’s important to note that all the studies were retrospective in design, making them more susceptible to publication bias [37]. Sensitivity analysis showed the significant effect remained after removal of the study with some risk of bias concerns (OR 1.95, 95% CI 1.38-2.76). Evidence from the subgroup with unadjusted data was considered low certainty (due to a high risk of bias and imprecision) and resulted in inconclusive evidence.

## DISCUSSION

### Summary of results

To our knowledge, this systematic review is the first to examine the evidence on the association between delirium and falls in community settings. Eight studies, encompassing at least 3505 unique participants from five high-income countries, were predominantly conducted in residential care settings. Due to the difficulty in establishing the temporal association between delirium and falls we included evidence from studies both where delirium preceded falls (D-F) and where falls preceded delirium (F-D). We suggest it is likely that in studies where delirium preceded falls, the delirium may have presented too far in advance, up to 6-12 months in some studies, to impact the falls directly although there may be an ongoing longer-term effect on fall risk. Conversely, where the fall took place before the delirium it is also possible that the delirium was present when the fall took place and was subsequently recorded during follow-up period.

From the five D-F studies, the one study presenting adjusted data showed a prior diagnosis of delirium was associated with a significant increase in the number of falls. However, this was low certainty evidence. The four unadjusted studies showed no clear association although the directions of effect were consistent with the adjusted data.

Pooled evidence from the three F-D studies which accounted for confounding factors showed a significant association between delirium and an increased falls risk. These studies were high quality, but with the caveat that they were retrospective and therefore highly susceptible to a number of biases, discussed below. The single subgroup which presented unadjusted data showed an inconclusive association between falls and delirium risk, but a consistent direction of effect.

### Strengths and limitations

This systematic review has a number of strengths. We followed transparent and robust methods to summarise the available evidence for the association between delirium and falls in a community setting. We included studies both where delirium preceded documented falls, and where falls occurred before a recorded episode of delirium to ensure our review was as comprehensive as possible. Mirroring the approach for prognostic studies, we included observational studies: cohort, cross-sectional, and case-control design. We assessed the risk of bias using the appropriate Newcastle Ottawa scale, considering both the overall rating and the individual domain ratings to arrive at our judgements. Evidence was assessed for certainty using an approach based on GRADE for prognostic studies as a general guide (although GRADE assessments were not specifically undertaken) [27]. This was a review of association and cannot be considered a prognostic review due to limitations of the primary study evidence.

There are some limitations to this review and the included studies. We incorporated observational studies utilising retrospective data and relying on patient records. The clarity of the data collection and recording process was sometimes insufficient. Such studies are susceptible to recall bias, incomplete record-keeping, and potential inaccuracies [38–40]. Furthermore, they may also be influenced by selection biases such as self-selection, non-response, and attrition, areas largely overlooked in the discussions of the included studies. Finally, studies reporting associations are at high risk of publication bias, and findings might not be reported if there is no observed association between variables [32].

We found a predominance of supported care settings in the studies. The definition of residential / nursing homes varies in different countries and the level of support, health and care provision and independent living varied across studies. The lack of studies focusing on data from falls in people’s own homes may be due to the lower incidence and prevalence of recorded delirium and falls in these settings [9, 11]. Very large studies with substantial datasets would be needed to accurately establish any association between delirium and falls in the home setting, and many episodes of delirium and falls in the community are not accurately recorded in healthcare records.

A number of factors make the comparison of findings between studies less robust. A wide range of assessment tools were used to measure delirium; in some cases, it was not clear how delirium had been identified and assessed. No study included a measure of the severity of the delirium or explored how this impacted on the number of falls. Additionally, definitions of delirium and falls were not universally presented in the papers, and we also included studies with author’s own definition or no definition. Furthermore, none of the studies collected specific data to investigate the link between delirium and falls, further limiting the evidence’s directness.

Of the three F-D studies that presented adjusted data, two included dementia in their multivariate models and one included cognitive impairment (but not dementia) (see Supplementary S5 for a full list of variables considered in the analysis). This is important because the co-existence of dementia is a potential confounding factor in the development of delirium, and delirium a major factor in the development of dementia [41]. The studies had a follow-up period of between one and sixteen months. However, there is a lack of data on the extended, longer-term effects of the relationship between falls and delirium in a community setting.

None of the included studies in this review made any consideration of equity factors, as identified in the PROGRESS Plus framework [42] in their analysis and interpretation of the data. This is crucial because environmental and equity factors may affect both falls and delirium, and some populations may be at greater risk; these factors were outside the scope of the current review..

Finally, we only searched for peer-reviewed publications written in English. We may have missed unpublished studies, pre-prints, and studies in other languages. Together with the risk of publication bias this increases the risk that relevant studies may not be included in this review, that the pattern of missingness is non-random and that the strength of the associations reported here may be an overestimate of the true effects.

### Conclusions and further research

We found a limited amount of evidence for an association between a record of delirium and an increase in falls, and evidence for an association between falls and an increased risk of a recorded delirium episode. However, these findings are based on studies with relatively small numbers of participants, and we note the potential for publication bias impacting these types of studies. The limitations of the studies and data do not allow direct comparison between the relative risks of D-F and F-D but there is enough evidence to suggest an association (both unadjusted and adjusted).

Our review highlights the need for more methodologically rigorous research to quantify the relationship accounting for important potential confounders or mediators such as dementia, frailty and polypharmacy. We recommend the use of standardised definitions and assessment measures for delirium and falls, e.g., CAM [43] or 4AT [44] for delirium and the ProFaNE core outcome dataset for falls research [1]. Additionally, clear identification of the timing of delirium and falls, specifically determining which occurred first, and more studies focusing explicitly on the association between delirium and falls would strengthen the body of high-certainty evidence.

Further research is needed to understand the complex relationship between delirium and falls, establish how and why this operates bidirectionally and identify the potential modifying factors involved. Recognition and focus on equity factors in studies may also help to identify groups more at risk of delirium and falls and allow for tailored interventions to meet the needs of underserved populations. It is imperative that studies finding null results are published.

This work has implications for clinical practice and policy including the implementation of routine screening for both delirium and falls to identify and treat preventable fall risks. In hospital settings patients are screened using 4AT for delirium and assessed for fall risk. We suggest the 4AT may also be suitable for rapid screening in the community setting, building on ongoing development of a community delirium toolkit [19, 45]. Enhanced screening and diagnosis may yield benefits, including

a. early delirium detection and treatment; b) reduced risk of subsequent falls; c) reduction in complications of delirium or falls; and d) reduced need for hospital transfer or inpatient care. Appropriately developed enhanced pathways considering the potential delirium-falls link may reduce personal and financial burdens to the individual and healthcare system Clinicians should be alert to the potential relationship between the two.

## Data Availability

This is a review of previously published studies, therefore all the data used in the review are already in the public domain. All data relevant to the included studies are included in the article or uploaded as supplementary information

## Notes

### Competing Interest Statement

The authors have declared no competing interest.

### Clinical Protocols

https://www.crd.york.ac.uk/prospero/display_record.php?RecordID=309982

### Funding Statement

CET, CS, SA, AM and CT are funded / partially funded by the National Institute for Health Research Applied Research Collaboration Greater Manchester (grant number: NIHR200174).

