## Supplementary files for "The association between delirium and falls in older adults in the community: a systematic review"

**Index**

**Supplementary S1: Example search strategy Medline (Ovid)**

**Supplementary S2: Inclusion and exclusion criteria**

**Supplementary S3: Additional study outcomes**

**Supplementary S4: 4a. Risk of Bias for studies with delirium preceding falls and 4b. Risk of bias for studies with falls preceding delirium**

**Supplementary S5: Variables accounted for in adjusted studies**

### Supplementary S1: Example search strategy Medline (Ovid)

Ovid MEDLINE(R) <1946 to April Week 4 2023>

| 1 | exp Delirium/ |
| --- | --- |
| 2 | deliri*.mp. |
| 3 | acute psychosis.mp. |
| 4 | acute psychoses.mp. |
| 5 | exp Psychotic Disorders/ |
| 6 | psychoses.mp. |
| 7 | exogenous psycho*.mp. |
| 8 | toxic psycho*.mp. |
| 9 | ICU syndrome.mp. |
| 10 | ICU psychosis.mp. |
| 11 | ICU psychoses.mp. |
| 12 | acute confusional state.mp. |
| 13 | acute confusional syndrome.mp. |
| 14 | toxic confusion.mp. |
| 15 | acute confusion.mp. or exp Confusion/ |
| 16 | temporary confusion.mp. |
| 17 | acute organic psychosyndrome.mp. |
| 18 | acute psycho-organic syndrome.mp. |
| 19 | acute brain dysfunction.mp. |
| 20 | acute brain failure.mp. |
| 21 | acute brain syndrome.mp. |
| 22 | organic mental disorder*.mp. |
| 23 | metabolic encephalopathy.mp. or exp Brain Diseases, Metabolic/ |
| 24 | exp Cognitive Dysfunction/ or exp Cognition Disorders/ or exp Memory Disorders/ |
| 25 | Neurocognitive Disorders/ |
| 26 | clouded state.mp. |
| 27 | clouding consciousness.mp. |
| 28 | 1 or 2 or 3 or 4 or 5 or 6 or 7 or 8 or 9 or 10 or 11 or 12 or 13 or 14 or 15 or 16 or 17 or 18 or 19 or 20 or 21 or 22 or 23 or 24 or 25 or 26 or 27 |
| 29 | exp Accidental Falls/ |
| 30 | fall.mp. |
| 31 | falling.mp. |
| 32 | fell.mp. |
| 33 | fallen.mp. |
| 34 | faller.mp. |
| 35 | fall injury.mp. |
| 36 | (slip and fall).mp. |
| 37 | (fall and slip).mp. |
| 38 | slipping.mp. |
| 39 | slip.mp. |
| 40 | trip.mp. |
| 41 | fall risk.mp. |
| 42 | fall-related.mp. |
| 43 | fall prevention.mp. |
| 44 | 29 or 30 or 31 or 32 or 33 or 34 or 35 or 36 or 37 or 38 or 39 or 40 or 41 or 42 or 43 |
| 45 | community.mp. or exp Residence Characteristics/ |
| 46 | community-dwelling.mp. or exp Independent Living/ |
| 47 | dwell* in the community.mp. |
| 48 | liv* independently.mp. |
| 49 | ageing in place.mp. |
| 50 | aging in place.mp. |
| 51 | nursing home.mp. or Home Nursing/ |
| 52 | care home.mp. or Home Care Services/ |
| 53 | exp Residential Facilities/ or exp Homes for the Aged/ or residential home.mp. |
| 54 | exp Long-Term Care/ |
| 55 | 45 or 46 or 47 or 48 or 49 or 50 or 51 or 52 or 53 or 54 |
| 56 | 28 and 44 and 55 |
| 57 | exp Aged/ |
| 58 | exp Aging/ or ageing.mp. |
| 59 | elder*.mp. |
| 60 | older.mp. |
| 61 | older age.mp. |
| 62 | geriatric*.mp. |
| 63 | senior*.mp. |
| 64 | retire*.mp. |
| 65 | later life.mp. |
| 66 | later-life.mp. |
| 67 | late life.mp. |
| 68 | late-life.mp. |
| 69 | later lives.mp. |
| 70 | later-lives.mp. |
| 71 | exp Longevity/ |
| 72 | baby-boom*.mp. |
| 73 | post-world war.mp. |
| 74 | 57 or 58 or 59 or 60 or 61 or 62 or 63 or 64 or 65 or 66 or 67 or 68 or 69 or 70 or 71 or 72 or 73 |
| 75 | over 60.mp. |
| 76 | 60 and above.mp. |
| 77 | over sixty.mp. |
| 78 | over 65.mp. |
| 79 | 65 and above.mp. |
| 80 | over sixty five.mp. |
| 81 | 70 |
| 82 | sevent*.mp. |
| 83 | 75 or 76 or 77 or 78 or 79 or 80 or 81 or 82 |
| 84 | 74 or 83 |
| 85 | 56 and 84 |
| 86 | limit 85 to yr="1995 -Current" |

### Supplementary S2: Inclusion and exclusion criteria

|  | Inclusion | Exclusion |
| --- | --- | --- |
| Population | Adults, 60+ years old, worldwide:   - In the community (not limited to): home (including those with minimal dependence such as home help/ home care), sheltered accommodation, primary care/GP practices/surgeries, outpatient clinics, community/day centres, day/respite care and carer/community groups. Community settings also include care homes, residential homes, nursing homes and long-term care facilities. These studies will be treated as a subgroup for analysis, as they are more likely to have different baseline in terms of health, compared to other participant populations. - Adults with clinically stable pre-existing conditions and study recruitment must not be based on the presence of a specific condition. - People in disadvantaged, minority or vulnerable groups (not limited to): BAME, ethnicity, religion, low socioeconomic position, LGBTQ+, travellers and health inequalities experienced due to diversity and equality variations (adherence to PROGRESS Plus reporting). | - Hospital inpatients, people in hospices and long-term hospital. - People in prison. - Studies of children or adolescents (<18 years old). - People with identified/diagnosed end stage disease/ terminal conditions. - People with severe mental health conditions such as schizophrenia and psychosis - If studies report on mixed populations of eligible/ineligible populations and differentiation is not possible, they will be excluded. |
| **Variables** | - Adults identified as having both delirium and falls as outcomes will be included. - We are interested in delirium as a risk factor for falls, however fall followed by delirium will also be included, given confirmation of delirium prior to a fall may not be feasible in the community setting. | - Delirium related to alcohol withdrawal (delirium tremens), presence of alcohol or drug misuse will be excluded. |
| Context | - Community dwelling adults, aged 60+ years old, irrespective of geographic location. | - Not in defined age group. - Not in the community. - Participants recruited on the basis of a specific diseases or illness. - Only protocol provided. - Only cost data. - No outcomes reported - Not in the English language or with no suitable English translations available. |
| Outcomes | - Number of falls (self-reported, observational/medical notes, use of technology e.g. fall alarms, fall detectors, fall mats) – reported as incidence - Number of fallers/non-fallers/frequent fallers (individuals who sustained more than one fall) – reported as prevalence - Falls rate per person/year of follow up - Time to first fall - Number of injurious falls (categorised as; a) serious injury – fractures or injuries requiring inpatient treatment, b) moderate injury – wound bruises, sprains and cuts requiring treatment, c) minor injury – minor bruises or abrasions not requiring treatment; reduction in physical activity (due to pain, fear of falling) for at least three days, and d) no injury – no physical injury detected) - Number of delirium cases detected (observational/medical notes and/or validated tools) - Number of delirium episodes recorded (observational/ medical notes and/or validated tools) - Duration, severity and type of delirium (observational/medical notes and/or validated tools) |  |

### Supplementary S3: Additional study outcomes

| **Delirium-falls studies** | | | | | | | | | | | |
| --- | --- | --- | --- | --- | --- | --- | --- | --- | --- | --- | --- |
|  | **Incidence of falls** | **Prevalence of falls** | **Falls rate per person/ year of follow-up** | **Time to first fall** | **Multiple falls (people who fell >once** | **Incidence of injurious falls** | **Prevalence of injurious falls** | **Incidence of delirium cases detected** | **Prevalence of delirium** | **Number of delirium episodes** | **Duration, severity, and type of delirium** |
| Eriksson et al (2007) | n=69 | n=34 (41%) | 1.8 falls/person year (1.5 for men and 1.9 for women) | *n/r* | 52 falls from 17 people | *n/r* | Fallers sustaining fracture n=2 (2.9%) | *n/r* | n=7 (8.6%) | *n/r* | *n/r* |
| Kallin et al (2002) | n=163 | n=52 (62.6%) | 2.29 falls/person-year | *n/r* | 145 falls from 34 people | *n/r* | All fallers n=28 (53.8%)  5/18 (28%) single fallers  23/34 (68%) recurrent fallers | *n/r* | Total n=26 (31.3%)  Fallers n=19 (36.5%),  Non-fallers n=7 (22.6%). | *n/r* | n/r |
| Kallin et al, 2004 | n= 482 | n= 113 (56.8%) | *n/r* | *n/r* | n/r | n=154 (32%)  (n=17 delirium) | n=74 (65%) | n=48 (10%) | Total n=63 (31.7%)  Fallers n= 20 (31.1%) | *n/r* | *n/r* |
| Mahoney et al. (2000) | *n/r* | 1^st^ mth p.d.  n=46 (14.8%)  2^nd^ mth p.d. n=22 (7%)  3^rd^ mth p.d. n=17 (5.4%) | 2-week p.d.  8 falls/ 1000 person days  8-week p.d.  2.5 falls / 1000 person days  3 mth p.d.  1.7 falls / 1000 person-days | *n/r* | At 1-month post-discharge  n=33 (1 fall)  n=9  (2 falls)  n=4  (3+ falls) | 1^st^ month p.d. 11% of falls resulted in hospitalisation | *n/r* | *n/r* | Probable delirium = n=5 (1.6%)  Rate ratio 4.11 (95% CI) (0.85-19.79) | *n/r* | *n/r* |
| von Heideken Wagert et al. (2009) | Total falls n=304  (OH n= 118,  IH n = 186) | Total n=88 (40%) (OH n=35 (32%), IH n= 56 (50%)) | 2.17 falls/PY (OH = 1.42 falls/PY, IH = 3.25 falls/PY) | 71 ± 56 days (range 1-175 days) | *n/r* | Injurious falls/PY = 0.83%  Minor injury = 0.62%  Major injury = 0.21%  Falls resulting in fractures = 0.15%  People injured n=5 (26%) | *n/r* | *n/r* | Total n=44 (20%) (fallers n=24 (26%), non-fallers n=20 (16%)) | *n/r* | *n/r* |
| Falls-Delirium studies | | | | | | | | | | | |
|  | Incidence of falls | Prevalence of falls | Falls rate per person/ year of follow-up | Time to first fall | Multiple falls (people who fell >once | Incidence of injurious falls | Prevalence of injurious falls | Incidence of delirium cases detected | Prevalence of delirium | Number of delirium episodes | Duration, severity, and type of delirium |
| Boorsma et al. (2012) Nursing home subgroup | *n/r* | 21 (28.4%) | *n/r* | *n/r* | n/r | n/r | n/r | 8.90% | 20.7 / 100 person-years | *n/r* | *n/r* |
| Boorsma et al. (2012) Residential subgroup | *n/r* | 34 (30.4%) | *n/r* | *n/r* | n/r | n/r | n/r | 8.20% | 14.6 / 100 person-years | *n/r* | *n/r* |
| Perez-Ros et al. (2019) | *n/r* | 37 (44.6%) | *n/r* | *n/r* | n/r | n/r | n/r | 121 episodes | n=83 (18.7%) | 26.5% (n=22) two or more | *n/r* |
| Sabbe et al. (2021) | *n/r* | 24 (50%) | *n/r* | *n/r* | n/r | n/r | n/r | *n/r* | n=48 (14.2%) | *n/r* | *n/r* |
| *Glossary: p.d. = post discharge, OH = Ordinary housing, IH = Institutional housing, PY = person year, n/r = not reported* | | | | | | | | | | | |

### Supplementary S4: 4a. Risk of Bias for studies with delirium preceding falls and 4b. Risk of bias for studies with falls preceding delirium

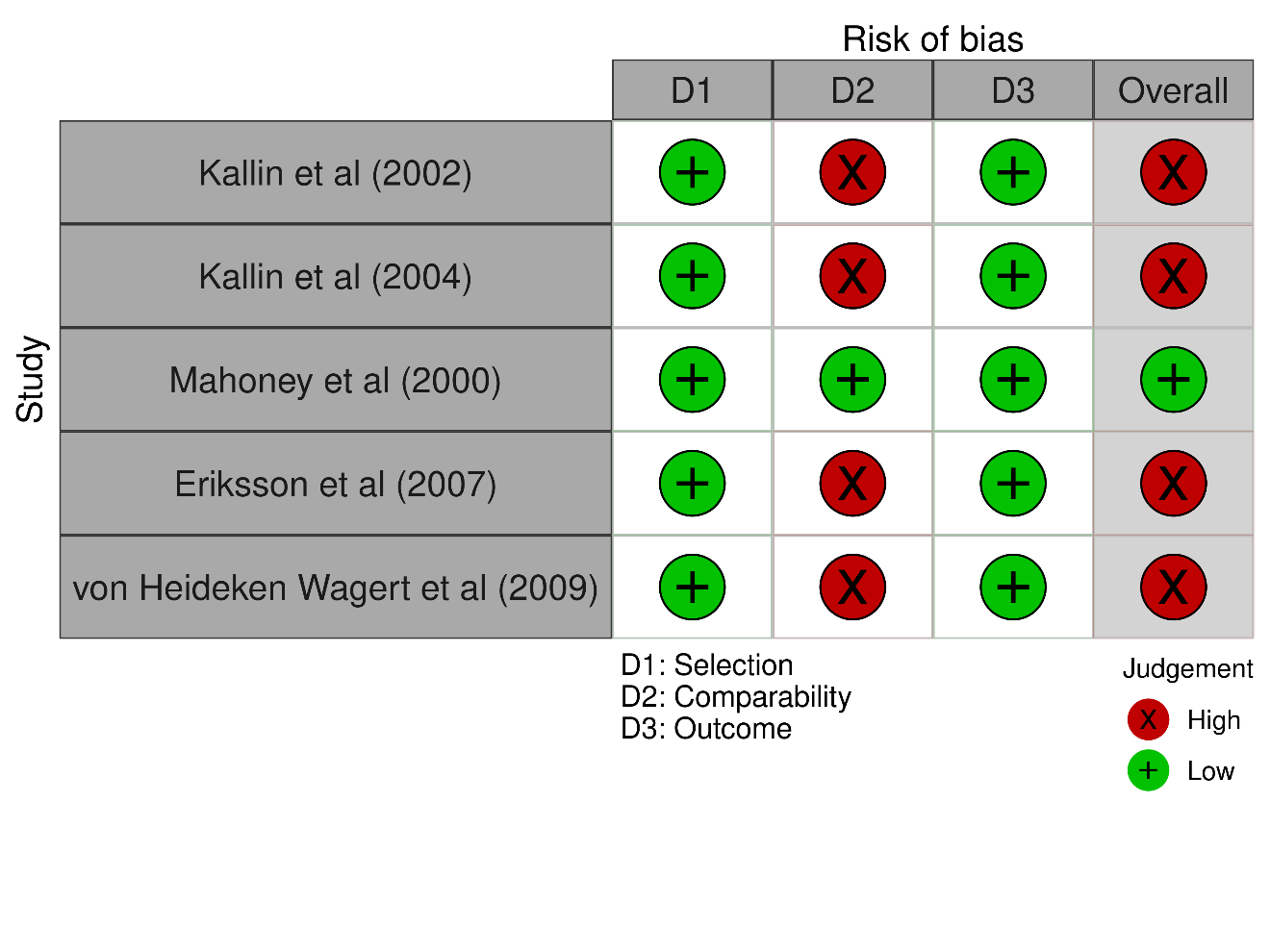

4a.

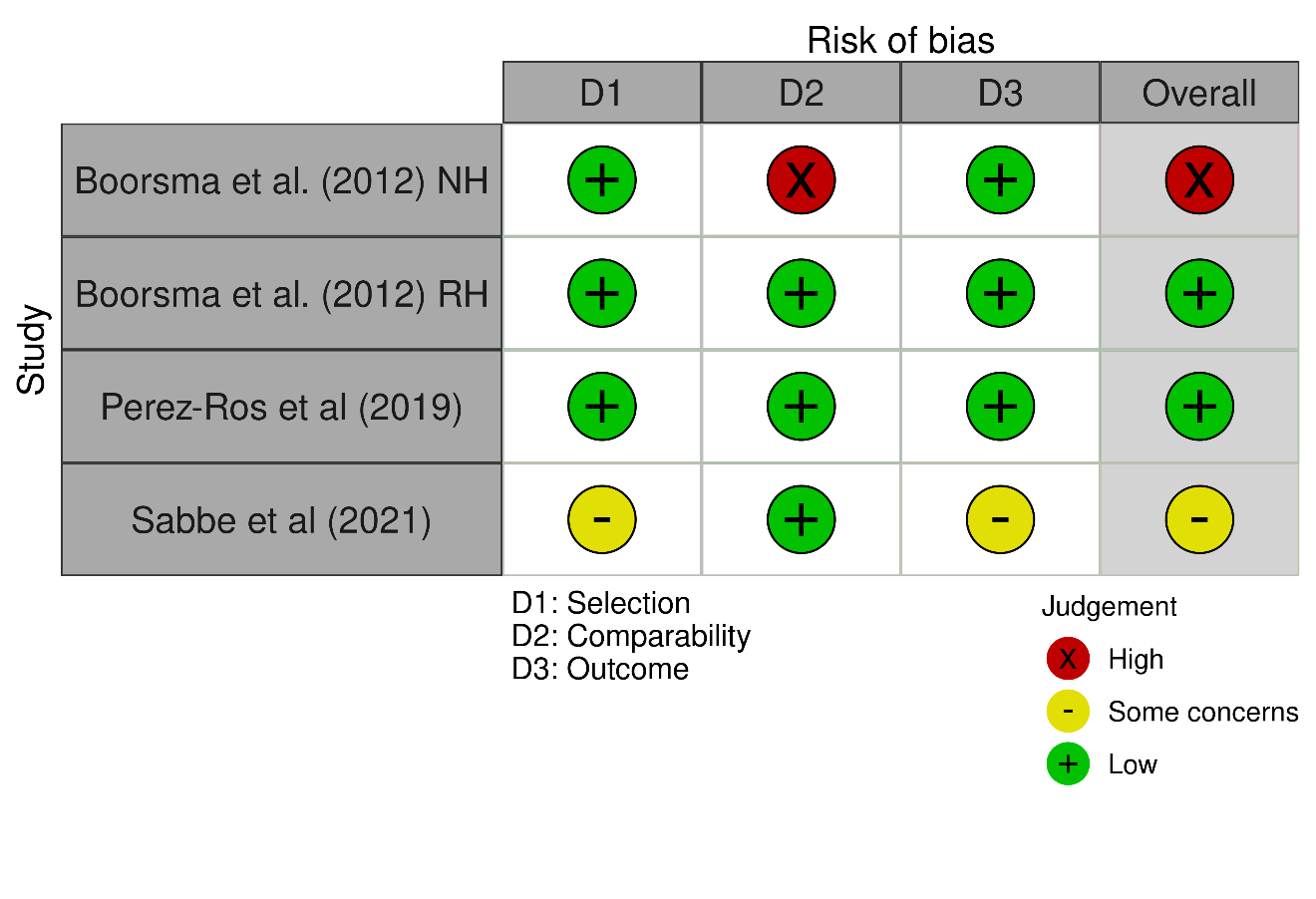

4b.

D1: 0-1 (high), 2 (some concerns), 3+ (low); D2: 0 (high), 1 (some concerns), 2+ (low); D3: 0 (high), 1 (some concerns), 2+ (low)

*Key: NH=nursing homes RH=residential homes*

### Supplementary S5: Variables accounted for in adjusted studies

| Adjusted data | Factors considered for adjusted models | Significant factors in model |
| --- | --- | --- |
| Delirium-falls |  |  |
| Mahoney et al 2000 | Prehospital  ≤2 wk prior  Dependent in ≥1 ADLs  Used ambulation aid indoors  Cane  Standard walker  None  Could not walk one block  Was light-headed with standing  Drank alcohol daily  Had a corrected distant visual acuity of  20/50 or worse  ≤1 y prior  Had ≥2 falls  No. of prior hospitalizations  Hospital  Admitting diagnosis  Cardiovascular  Pulmonary  Gastrointestinal tract  Neurologic  Other  No. of diagnoses at admission  Length of stay  Posthospital  No. of prescribed medications  No. of new medications  No. of psychoactive medications  Medication class prescribed  Cardiovascular  Long-acting benzodiazepine  Short-acting benzodiazepine  Tertiary amine tricyclic  antidepressant  Other antidepressant  Antipsychotic  Dependent in ADLs  Uses ambulation aid indoors  Cane  Standard walker  None  Cannot walk 1 block  Balance tertile score  Lowest (0-18)  Middle (19-22)  Highest (23-26)  Mini-Mental State Examination score  (maximum = 30)  Probable delirium  Feels sad or depressed  Orthostatic systolic BP decrease | Prehospital  Dependent in ≥1 ADLs  Used standard walker indoors  Had ≥2 falls in year prior  No. of hospitalizations in year prior\  Hospital  Admission  Gastrointestinal tract diagnosis  Posthospital  Tertiary amine tricyclic  antidepressant  Uses cane indoors  Balance score  Middle tertile  Lowest tertile  Probable delirium |

| Adjusted data | Factors considered for adjusted models | Significant factors in model |
| --- | --- | --- |
| Falls-delirium |  |  |
| Boorsma et al 2011 Residential Homes | Socio-demographics  Male  Older age  Chronic diseases  Dementia  Depression  Parkinson  Cardiovascular diseases  Diabetes  Care-related variables  Bed rails  Chair restraints  Trunk restraints  Psychological variables  Anxiety  Use of antipsychotics  Functional variables  ADL dependency  Infection  Visual impairment  Hearing impairment  Pain  Fall incidents  Fractures  Daily incontinence of urine  Bedridden | Dementia  Falls incidents |
| Perez-Ros et al 2019 | Delirium predisposing factors  Age ≥89 y  Male sex  Barthel score <60 points  Yesavage scale >5 points  MMSE <24 points  Diagnosed depression  Cornell >8 points  Diabetes mellitus  Stroke  Insufficient hydration  Epilepsy  Dementia  Comorbidity  Parkinson’s disease  Dysphagia  Hearing impairment  Visual impairment  Previous delirium  Delirium triggering factors  Polypharmacy (>7 drugs)  Anxiolytics  Anticholinergic agents  Antidepressants  Neuroleptics  Anemia  Emergency visits in previous 12 months ago  Autoimmune diseases  Falls  Glomerular filtration rate <60 ml/min  Hospital admissions in previous 12 months  Hypoglycemia in previous month  Incontinence  Renal failure  Pneumonia  Pain  Systemic infection (non-urinary)  Bladder catheter  Urinary infection | Dementia  Neuroleptics  Anticholinergic agents  Falls |
| Sabbe et al 2021 | Being physically restrained  Participate in activities  Never Baseline  Daily  Weekly  Two-weekly  Monthly  Fall incident < 90 days  Cognitive impairment  ADL score  Medication  Antipsychotics  Chronic pathology  Dementia  Pneumonia  Parkinson | Fall incident < 90 days  Cognitive impairment |
